# Plasma brain-derived tau: analytical and clinical validation of the first commercial immunoassay

**DOI:** 10.1101/2025.07.21.25331193

**Authors:** Michel N. Nafash, Sarah E. Svirsky, Xuemei Zeng, Yijun Chen, Julia K. Kofler, Ann D. Cohen, David O. Okonkwo, Oscar L. Lopez, Ava M. Puccio, Thomas K. Karikari

## Abstract

**Background:** Brain-derived tau (BD-tau) is a promising blood-based biomarker for neurodegeneration/brain injury in neurodegenerative and acute neurological disorders. However, widespread use is hampered by lack of commercial assays.

**Methods:** Using the Simoa® HD-X analyzer, we evaluated the first commercial research-use only BD-tau Advantage PLUS assay’s robustness, precision, dilution linearity, spike recovery, specificity, and limits of detection. Matrix effect was examined by comparing BD-tau levels in n=48 plasma/serum and n=20 plasma/CSF sample pairs. Clinical performance was examined in a traumatic brain injury (TBI) cohort.

**Results:** Twenty repeated measurements of three plasma samples gave intra- and inter-plate CVs ≤7.24%. A median drift of 8.00% (decrease) was observed from the start to the end of a full plate run. Analytically, BD-tau concentrations decreased linearly up to 16-fold dilution, spike recovery was 86-96%, and signals were highly specific to the CNS-abundant tau441 but not the peripherally-enriched “big-tau” isoform. Moreover, signals were stable for up to four freeze/thaw cycles. Furthermore, significant correlations were observed in the plasma/serum (r=0.8392; p<0.0001) and plasma/CSF (r=0.6150; p=0.0039) pairs. Finally, plasma BD-tau was elevated in severe-acute TBI vs. chronic-mixed TBI and unaffected controls (p<0.0001; AUC=0.9986, and p<0.0001; AUC=1.000, respectively). In severe-acute TBI patients, plasma BD-tau was correlated with plasma p-tau217 (r=0.5761, p=0.0005), NfL (r=0.8910, p=0.0001), and GFAP (r=0.5424, p=0.0011). CSF BD-tau and CSF p-tau217 were strongly correlated (r=0.9667, p=0.0002).

**Conclusion:** BD-tau Advantage PLUS produces robust brain-derived tau-specific readings that demonstrate utility in detecting severe-acute TBI.

## Introduction

Clinical care for neurological disorders is experiencing a shift towards the integration of biomarkers into standard practices for diagnosis, tracking disease progression, and measuring response to therapies [1]. A biomarker can be objectively quantified and, when thoroughly validated, can act as a surrogate indicator of disease state and stage, as well as target engagement and efficacy in therapeutic campaigns [1,2]. For example, in several neurodegenerative diseases, cerebrospinal fluid (CSF) biomarkers have been incorporated into clinical practice for over a decade in conditions like Alzheimer’s disease (AD) and Parkinson disease (PD) [3–8]. However, the utility of CSF biomarkers is limited by accessibility, invasiveness, and cost concerns [9,10]. Blood-based biomarkers have seen a fast emergence due to the ease of collection, affordability, and comparable performance to CSF biomarkers [4,8,9,11,12].

A main limitation of blood-based biomarker assays for neurological disorders is the potential contamination of blood measurements by protein forms in systemic circulation originating from peripheral sources and not released from the central nervous system (CNS) [6,13–16]. This issue affects the reliability of several prominent biomarkers when measured in plasma including amyloid-beta (Aß) peptides, given their high levels in both CNS and non-CNS sources such as platelets and the liver which compromises the fold change in plasma when compared with levels in CSF [13]. A similar situation exists for tau protein which in addition to being abundant in the CNS, is equally found in many non-CNS tissues including lungs, heart, kidneys, muscles, and the peripheral nervous system [6,14–16]. This accounts, at least partly, for why the neurodegeneration/brain injury biomarker total tau performs poorly in blood despite its high accuracies in CSF, which show no correlations to blood-based measurements in AD [6,17]. Therefore, optimization of blood-based biomarker assays to improve their selectivity for CNS-originating proteins is needed to improve their accuracies and alignment with their CSF biomarker assay counterparts while maintaining the integrity of their measurements.

In 2023, a blood-based assay referred to as brain-derived tau (BD-tau), which provides a “barcoding” approach to select for tau protein released from the CNS, was reported [6]. The main unique feature is that BD-tau uses a novel antibody combination that ensures signal specificity to analytes only containing the six tau isoforms abundantly found in the adult human brain vs. the higher molecular weight tau isoforms (containing the exon 4a insert) more commonly released from the periphery (thus the name big-tau) [6]. Plasma/serum BD-tau levels strongly correlated with CSF BD-tau and CSF total tau, outperforming blood-based total-tau in this regard [6,18]. Moreover, plasma and serum BD-tau levels are strongly correlated [18].

In addition to showing strong correlations with other plasma and CSF biomarkers of AD as well as with baseline and worsening cognitive decline, BD-tau levels were more related to neurodegeneration in AD vs. non-AD neurodegenerative diseases, and appeared unaffected by common comorbidities (e.g., diabetes) and *APOE ε4* carriership [6,19,20]. In addition, plasma/serum BD-tau levels agree strongly with the severity of neurodegeneration/brain injury have demonstrated clinical utility for rapidly progressive AD and Creutzfeldt-Jakob disease [21]. In severe traumatic brain injury (sTBI), initial BD-tau measurements at the time of and one week after injury were more robustly correlated with worse clinical outcome scores than plasma total-tau, p-tau231, NfL, showing the significance of CNS selective-tau in outcome prediction [22].

Finally, BD-tau shows a more coherent time-dependent pattern to predict outcome of traumatic brain injury than both NfL and total-tau [22].

To date, BD-tau studies have been limited to using the original prototype assay. More recently, a commercial version of the blood-based BD-tau measurement assay, the Quanterix® BD-tau Advantage PLUS reagent kit, became available. However, independent analytical and clinical validation is lacking. In this study, we performed analytical validation tests according to the FDA’s recommendations for bioanalytical method validation [23], as well as analytical chemistry experts, on the aforementioned assay to ensure the accuracy and reliability of measurements [24]. We also report validation of the assay in clinical severe TBI patients.

## Materials and Methods

### Study Cohorts and Sample Collection

Plasma samples used for validation experiments were collected from participants at the University of Pittsburgh’s Alzheimer’s Disease Research Center (ADRC) into ethylenediaminetetraacetic acid (EDTA) tubes. Samples were spun at 2000xg for 10 minutes at 4°C to separate plasma from whole blood, then aliquoted into microtubes and stored at -80°C until time of use. Storage conditions for samples collected purposefully for assay stability experiments differed from this protocol as highlighted in Supplemental Table 1. Additionally, a pooled plasma sample commercially acquired from Precision for Medicine Bethesda, MD, USA was aliquoted into 500µL single-use aliquots and stored at -80°C until needed. Plasma and serum pairs were collected from the ADRC cohort following procedures described previously [25] and stored at -80°C pending use. Biobanking measures followed standard guidelines [26].

Severe TBI participants were prospectively enrolled at admission between 2019-2024 through the Brain-Trauma Research Center at the University of Pittsburgh, Pittsburgh, PA, USA. Eligibility was determined by the presence of severe TBI defined as Glasgow Coma Scale (GCS) score <9 at enrollment and age 16–80 years. Exclusion criteria were penetrating brain injury, brain death and pregnancy.

Chronic mixed TBI and control individuals were recruited through a Peer Reviewed Alzheimer’s Research Program (PRARP) study. Inclusion criteria for the chronic mixed TBI group were: 1. History of at least one TBI that occurred over one year prior to enrollment; 2. Age 29-59 years; 3. Suspected cognitive impairment (based on neuropsychological assessment or professional diagnosis of cognitive impairment). Exclusion criteria for the chronic mixed TBI group were: 1. History of penetrating TBI, 2. history of pre-existing neurologic or neurodegenerative disorder, 3. History of psychiatric disorder, 4. Active drug or alcohol dependence, 5. Contraindication to magnetic resonance imaging or PET scans. Inclusion criteria for the control group were age 29-59 years and a score less than 1 the Ascertain Demential 8 item Questionnaire (AD8), and exclusion criteria were a history of blast exposure or moderate to severe TBI.

Blood was taken from an indwelling catheter placed as a routine standard of care using 6 mL K3EDTA tubes (Greiner Bio-One, Fischer Scientific). To isolate plasma, samples were spun for 15 mins at 1500xg within 60 minutes of collection and aliquoted into cryovials for storage at -80 °C. For the severe TBI group plasma collected on day 4 post-injury was used in this study. For the chronic mixed TBI and control groups, baseline plasma was collected at time of consent, using similar methods.

A subset of the chronic mixed TBI (n=7) and control subjects (n=7) had CSF collected by lumbar puncture. CSF was collected from sTBI participants (n=9) using an indwelling ventriculostomy catheter via external ventricular drainage. All samples were gently inverted to avoid possible gradient effects, briefly centrifuged at low speed, and aliquoted into polypropylene cryovials prior to freezing and storage at –80°C.

The Pittsburgh ADRC and the Brain-Trauma and PRARP TBI studies were reviewed and approved by the Institutional Review Board at the University of Pittsburgh. The reference numbers for approvals are as follows: ADRC study, MOD19110245-023; BTRC study, 19030228; Military CTE study, 19110161.

### Data Collection

Data for this study were collected and analyzed at the University of Pittsburgh’s Biofluid Biomarker Laboratory between June 2024 and March 2025. Biomarker tests were run on Quanterix® Simoa^®^ HD-X analyzer using the Quanterix^®^ BD-tau Adv PLUS reagent kit (Lots 504274 and 504275). All average enzymes per bead (AEB) and replicate concentration values were acquired directly from instrument-generated exports, except for concentrations calculated with the custom-generated universal calibration curve. Calibrators were run in either duplicates or triplicates. We excluded calibrator H (60.0 pg/mL) after determining its non-importance for estimating BD-tau concentrations in plasma and serum based on interpolating concentrations with calibration curves using different subsets of calibrators.

### Experiment Procedures

Analytical validation experimental procedures are described in Supplemental Appendix 1.1.

### Statistical Analyses

All statistical analyses were performed in GraphPad Prism (v10.1.2(324)) and Microsoft Excel (V2503 Build 16.0.18623.20266). Descriptive statistics, Spearman correlations, Friedman and Dunn’s multiple comparisons tests, and Kruskal-Wallis one-way ANOVA and multiple comparison tests were conducted using GraphPad. Since data were not normally distributed, non-parametric tests were used and the results presented as median (IQR) values. Prism was also used for simple linear and 4-parameter logistic (4-PL) regression models, the latter being used to generate the universal calibration curve. Microsoft Excel was used to organize the preparation and results of tests such as dilution linearity and spike recovery, for normalization of the precision and sample stability tests, and to apply the 4-PL model to generate assay concentrations. For the lower limit of quantification (LLOQ) calculations, 4-PL parameters generated by the instrument were used, whereas the universal calibration curve parameters were acquired from GraphPad Prism’s interpolation of the calibrator AEB data. Finally, QC performance and calculation of the coefficients of variation (CV) were also done using MS Excel.

## Results

### Cohort characteristics

Characteristics of the participants (n=50) who provided the plasma/serum pairs have been described previously [25]. Characteristics of the participants (n=63) of the pilot TBI cohort are found in Supplemental Table 2.

### Assay details

We used an eight-point calibration curve for the BD-tau assay, with seven non-blank calibrators ranging from 0.246 pg/mL to 150 pg/mL. See Supplemental Figure 1 for a sample calibration curve.

### Robustness and Precision

We assessed the robustness, repeatability, and intermediate precision of the assay under two different conditions. First, three QC samples with different BD-tau levels were used to evaluate assay reproducibility. For samples analyzed on the same plate (n=10 independent runs), the median CV for the high QC sample [median: 223 pg/mL (IQR: 207-227)] was 5.65%, versus 5.23% and 7.03% for the medium and low QC samples [16.4 pg/mL (15.2-16.7)] and [7.42 pg/mL (7.07-7.59), respectively]. These %CVs were within the widely accepted range of <20% [24]. Furthermore, the median drift in signal from the start to the end of the same plate for aliquots of identical QC samples was ∼8.00% (range: 7.40-13.9%) [Figure 1A]. Overall assay robustness was good with a %CV of up to ∼7.24% for the pooled between-plate data [Figure 1B]. These results suggest that the assay is robust for independent measurements performed at different times.

**Figure 1.**
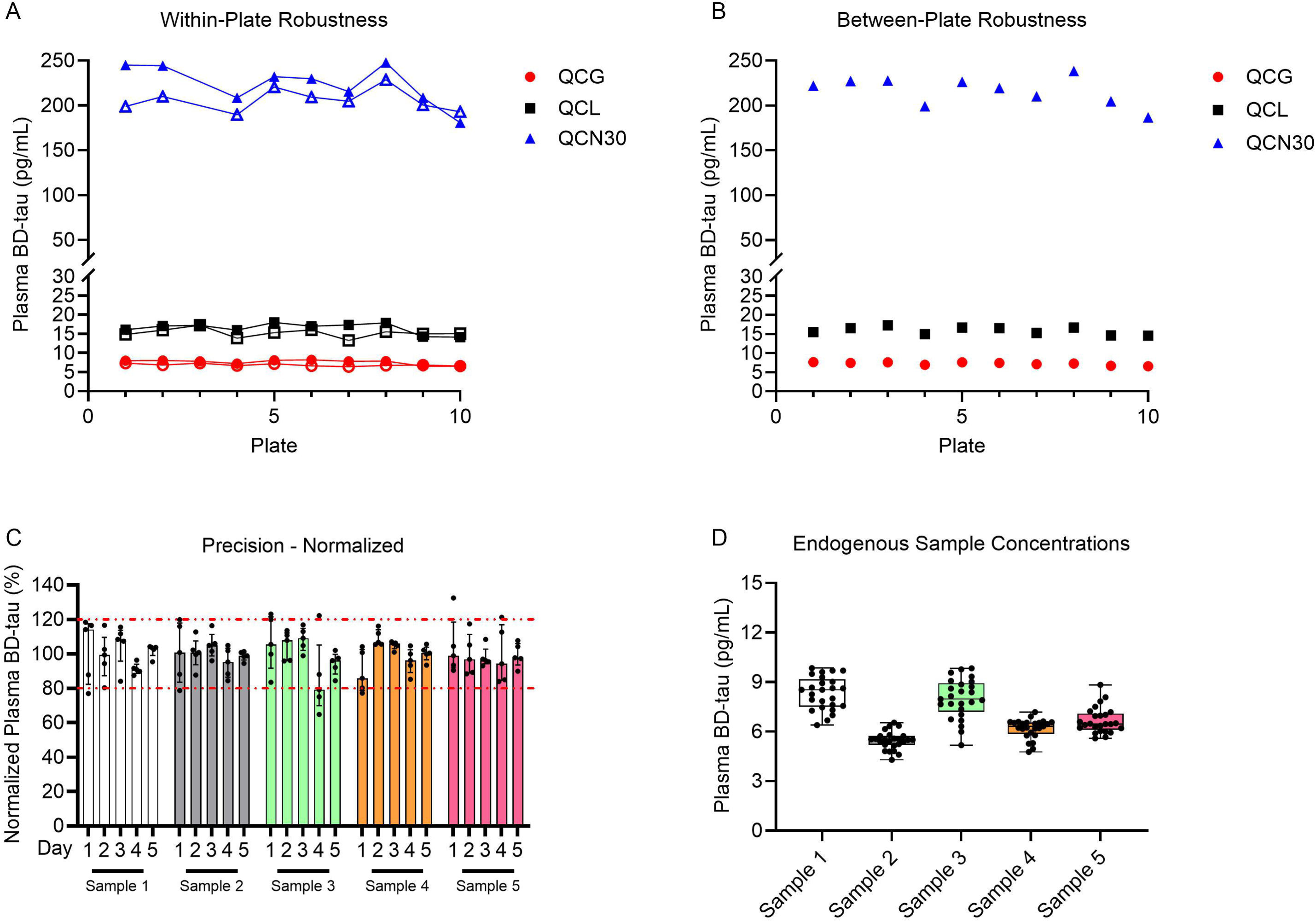
Robustness and precision of plasma BD-tau. (A) Plot showing pooled data across 10 independent runs for three different pre-made QC samples. Each sample was measured in duplicates at the start and at the end of the plate. The mean values at the start (solid) vs. the end (hollow) for readings from the same plate are shown. (B) Plot of the overall average readings for each QC per plate (combining the duplicate readings at the start and at the end). (C) Normalized plasma BD-tau concentrations for the five samples evaluated in the precision experiments. Single-use aliquots of each specimen were measured 25 individual times, across five runs over five different days. For each sample, the black dots show the individual concentrations in a single precision run, the bars show the median, and error bars show the inter-quartile range (IQR). (D) Measured (without normalization) sample concentrations in the precision experiments.

The second condition involved five different plasma samples that were measured 25 individual times, across five runs over five different days. For each sample evaluated, Figure 1C shows the % plasma BD-tau levels normalized to the mean of all 25 measurements, used to quantify repeatability and intermediate precision. The repeatability (CV_r_) was 6.47%-12.4% and the intermediate precision (CV_Rw_) 9.52%-15.1% [Supplemental Table 3]. The measured endogenous concentrations of each sample are shown in Figure 1D and Supplemental Table 2. This set of results indicates that the assay is robust, and that results can be compared to one another within the same batch/lot.

### Limits of Quantification

AEB signals for 16 blank samples ranged between 4.290x10^-3^ and 8.740x10^-3^ [Supplemental Table 4], with the mean and SD being 5.990x10^-3^ and 1.070 x10^-3^, respectively. The LLOQ signal (mean + 10 standard deviations) becomes 0.0167, which gives an analytical LLOQ of 0.0933 pg/mL when calculated according to Equation 1 [Supplemental Appendix 1.1.3]. The manufacturer’s analytical LLOQ is slightly higher at 0.133 pg/mL [27]. After adjusting for the 4X on-board dilution recommended for plasma samples, the functional LLOQ becomes 0.373 pg/mL.

For the ULOQ determination, the median concentration of the sample with the highest signals being 460 pg/mL, which gives a functional concentration of 1840 pg/mL after adjusting for the 4X on-board dilution. However, the highest point in the dynamic range of the assay calibration curve is 150 pg/mL (600 pg/mL dilution adjusted concentration) and marks the end of the dynamic range as published by the manufacturer [27]. Thus, 600 pg/mL is the highest point that can be reliably produced by the assay’s calibration curve.

### Signal specificity

We compared BD-tau results for equimolar concentrations of the recombinant CNS- abundant full-length tau441 (2N4R) isoform vs. the much longer big tau isoform commonly expressed in peripheral tissue in both calibrator diluent (manufacturer recommended) and four-fold diluted low-concentration QC plasma to account for potential matrix effects. Across three concentrations tested within the dynamic range of the assay (25 pg/ml, 50 pg/mL, and 100 pg/mL), signals for the 2N4R tau441 isoform increased proportionally to the dilution fold when diluted in either diluent or QC plasma. However, the big tau isoform signals remained similar to the blank values [Figure 2, Supplemental Table 5]. This indicates that the assay is highly specific for CNS-abundant tau isoforms.

**Figure 2.**
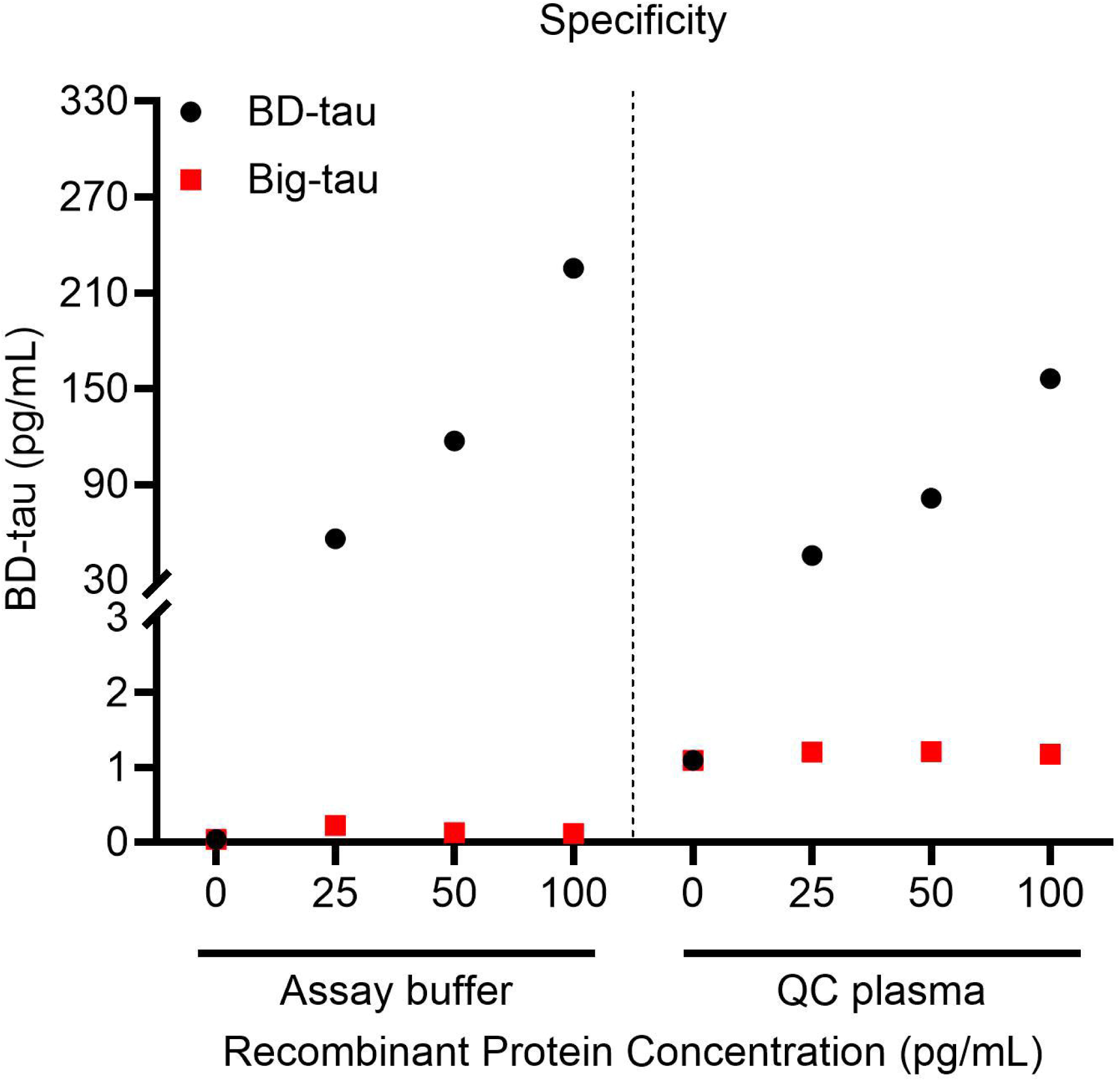
BD-tau assay specificity. Comparison of BD-tau assay readings when measured in equal concentrations of recombinant forms of 2N4R CNS-abundant tau441 and the peripherally-enriched big tau isoform. The test proteins were evaluated in two separate experimental conditions involving three theoretical concentrations of the recombinant protein and the non-spiked matrix for background signal: (1) dilution in the calibrator diluent following the manufacturer recommendations, and (2) four-fold dilution in QC plasma to account for potential matrix effects.

### Dilution Linearity

Dilution linearity data of three plasma samples were obtained from serial four-fold dilutions up to 4096-fold [Figure 3A]. Compared with the undiluted stock, the observed concentrations for all samples at the 4- and 16-fold dilutions were as expected, with measured values decreasing proportionally. The mean dilution-adjusted relative error was 112% for 4-fold dilution and 120% for 16-fold dilution [Figure 3B]. For the non-diluted, and 4- and 16-fold dilutions, the R^2^ was 0.908, 0.903, and 0.906 for plasma samples 1, 2, and 3, respectively.

**Figure 3.**
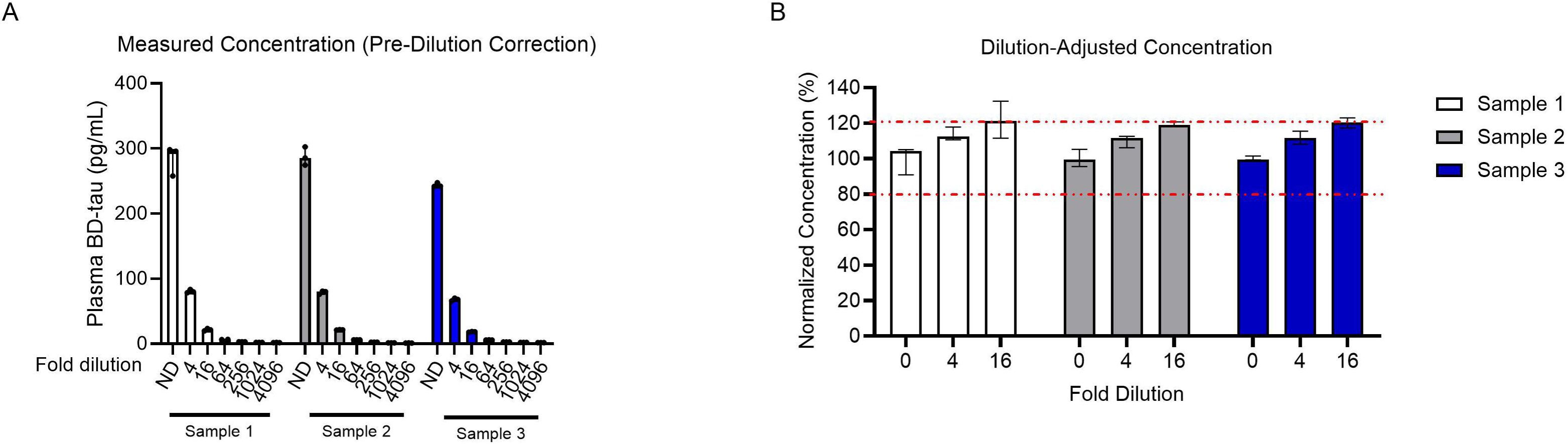
Dilution linearity of plasma BD-tau. A) Portrayal of dilution linearity of plasma samples showing the concentration from non-diluted (ND) up to 4096-fold diluted, across four-fold stepwise decreases. B) The dilution-adjusted relative errors at 4- and 16-fold dilutions for samples showing the adjustment which produced concentration within ±20% the normalized concentration of the dilution-free (neat) sample (shown as dotted red lines at 80% and 120%). Dilutions beyond 16-fold are not shown since their normalized concentration exceeded 140%.

However, after the 64X dilution the samples stopped responding to dilutions. The dilution-adjusted relative results error showed that beyond 16-fold dilution, the error becomes extremely high and can potentially lead to falsely high concentrations of plasma BD-tau. Together, the results show that the BD-tau assay concentrations change linearly with the degree of dilution up to 16-fold depending on the starting concentration.

### Spike Recovery

Spike recovery results were obtained from two different four-fold diluted plasma samples with non-spiked neat concentrations of 1.19 pg/mL and 1.67 pg/mL for Sample 1 and Sample 2, respectively. For both samples, recovery was within the 80-120% acceptable range for all chosen spike concentrations [Figure 4 and Supplemental Table 6].

**Figure 4.**
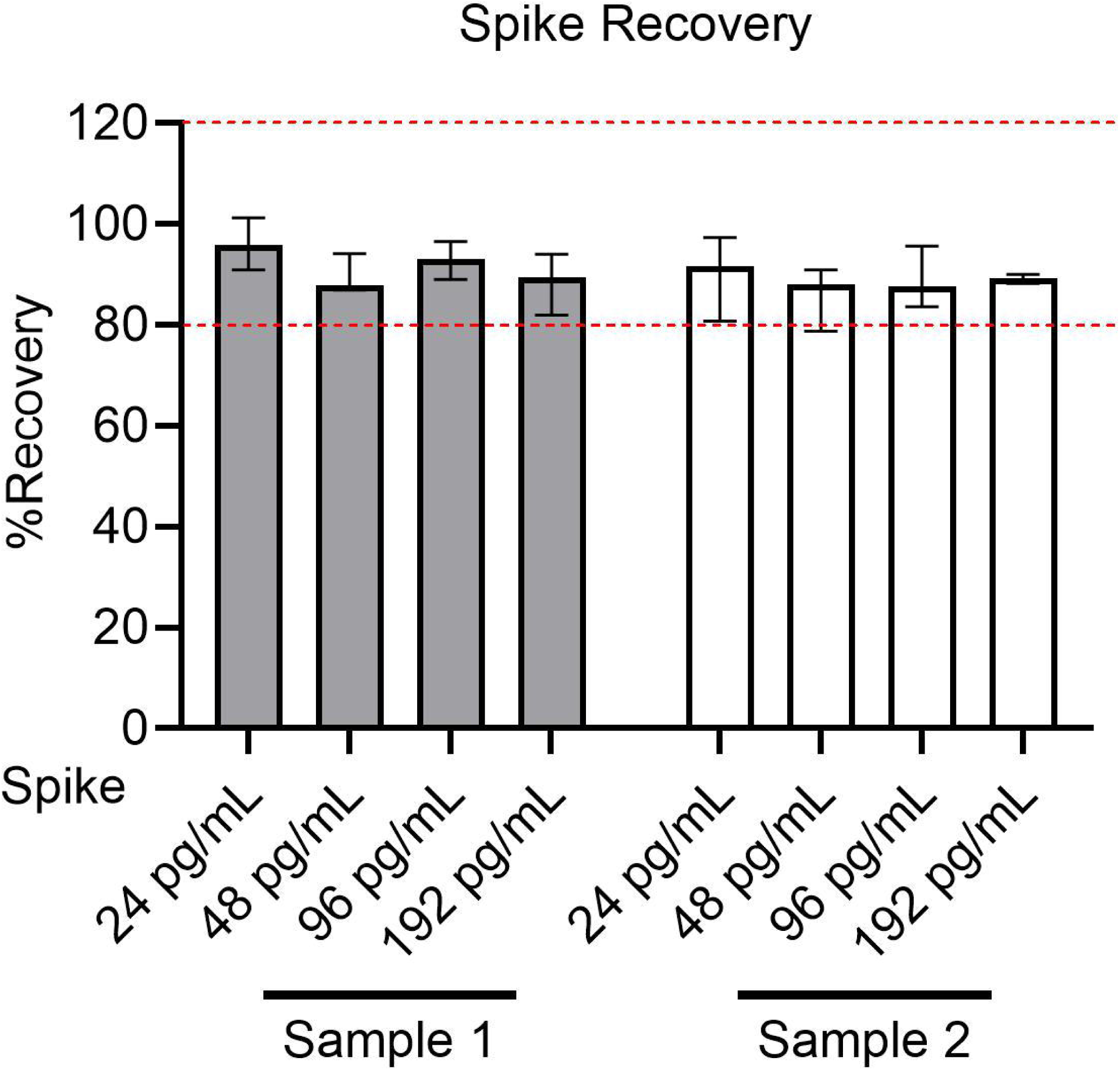
Spike recovery of plasma BD-tau. Spike recovery in two four-fold diluted plasma samples at four different spike levels. The bars indicate the percentage recovery in each sample at the given spike level, and the red dotted lines at 80% and 120% show the acceptable range of recovery as described by field experts *[24]*.

### Plasma/Serum Comparison

Of the n=50 plasma/serum pairs compared, 48 pairs gave values that were eligible for analysis, as two returned technical errors. Although BD-tau levels were higher in serum vs. in plasma [Figure 5A], there was a significant positive correlation (Spearman’s rho = 0.8392, p<0.0001) between the levels in the paired samples [Figure 5B].

**Figure 5.**
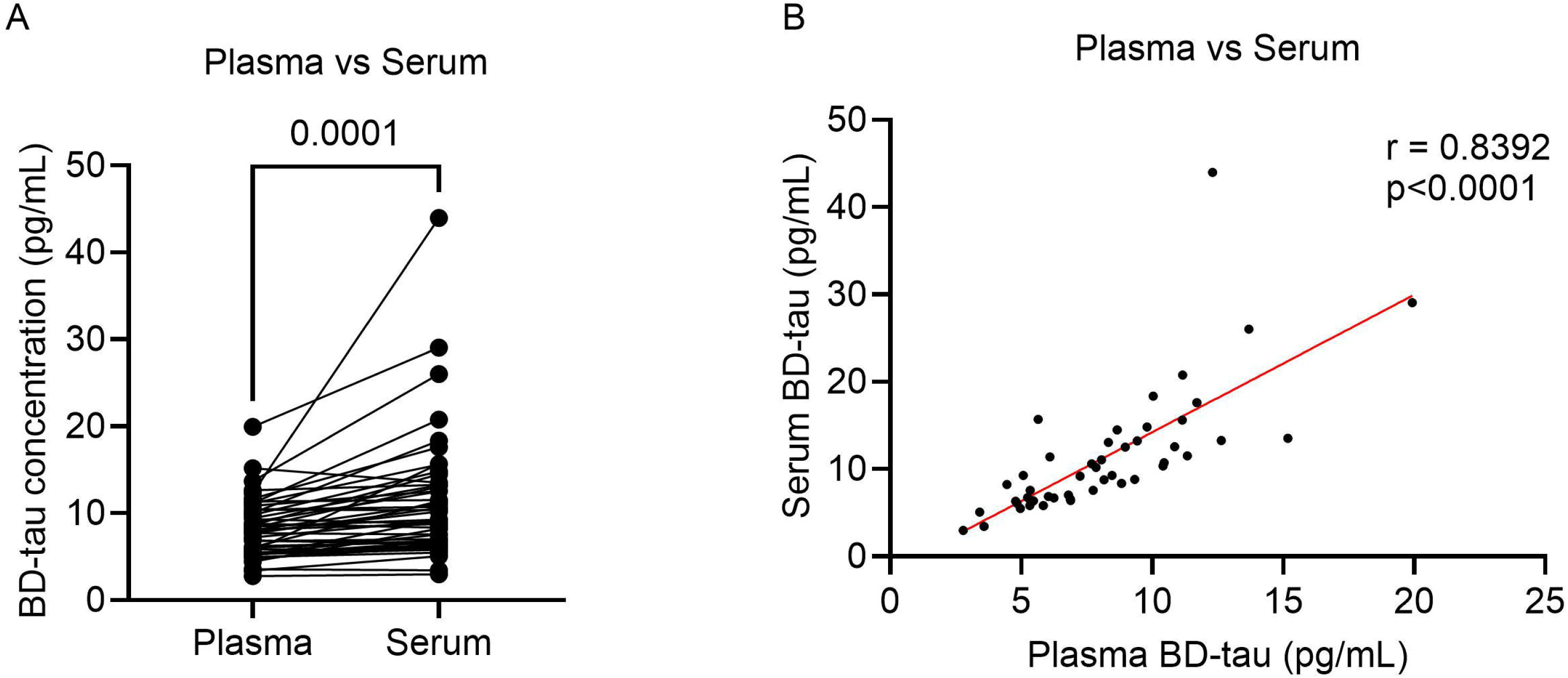
Comparison of BD-tau levels in paired plasma vs. serum samples. (A) Comparison of the absolute concentrations of BD-tau in paired plasma/serum samples from the University of Pittsburgh ADRC (n=48 viable pairs from a cohort of n=50). (B) Spearman correlation between plasma and serum BD-tau concentrations. Each point represents a plasma/serum pair, shown in red is the linear regression line.

### Sample Stability

Stability of three independent plasma samples was tested after storage in different temperature conditions and repeated freeze/thaw (f/t) cycles. Normalized concentrations showed that the storage conditions before moving the sample to -80°C had no impact [Supplemental Figure 2]. Concerning the impact of f/t cycles, increasing the number of f/t cycles tended to increase the levels of plasma BD-tau. Friedman test revealed significant differences in sample concentrations between the freeze thaw cycles (p=0.0004), with Dunn’s multiple comparisons test showing significant differences from the reference sample at f/t cycles number 6 (p=0.0101) and 8 (p=0.0011) [Supplemental Figure 3]. These findings show that plasma samples for BD-tau measurement should not be exposed to more than four f/t cycles.

### Clinical validation in TBI

In the pilot TBI cohort that included n=64 participants with plasma samples, including 34 sTBI participants whose plasma was collected at day 4 post injury, with a subset of 20 additionally having CSF samples [Supplemental Table 2], participants were grouped as unaffected controls, chronic-mixed TBI (cmTBI), and severe-acute TBI (sTBI). Nonparametric Kruskal-Wallis one-way ANOVA tests indicated significant differences among the three groups in both CSF (p=0.0168) and plasma (p<0.0001) BD-tau. Subsequent multiple comparisons test showed that The Kolmogorov-Smirnov test showed that CSF BD-tau distinguished between sTBI and other groups (sTBI vs cmTBI, p<0.0001; sTBI vs control, p<0.0001), but not between cmTBI and controls [Figure 6A]. On the other hand, multiple comparisons tests in CSF did not reveal any significant differences between controls and either sTBI or cmTBI. However, a significant difference exists between sTBI and cmTBI (p=0.0286) [Figure 6B]. Overall, the median BD-tau readings in CSF/plasma pairs were 378 pg/mL and 9.23 pg/mL in CSF and plasma, respectively, suggesting that ∼2.47% (1.62-3.35) of the BD-tau values in CSF are reflected in plasma for the same individual.

**Figure 6.**
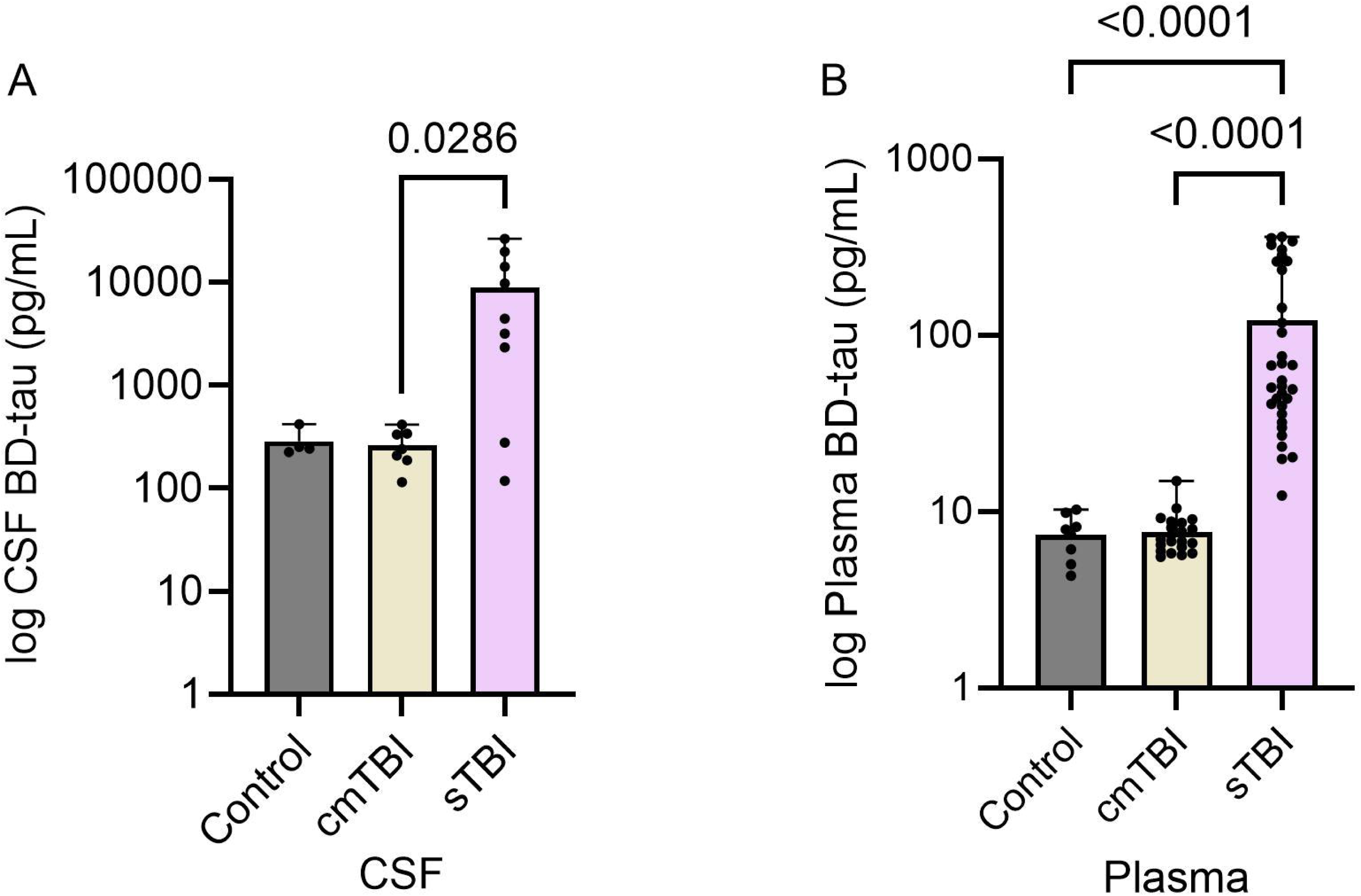
Clinical validation of plasma and CSF BD-tau in the pilot TBI cohort. BD-tau readings for control, chronic-mixed TBI (cmTBI), and severe-acute TBI (sTBI) participants from a pilot cohort in both CSF (A) and plasma (B), plotted on a log(10) scale. All data points are shown. Kruskal-Wallis multiple comparisons tests are shown comparing group BD-tau distributions where significant differences are seen.

Additionally, in TBI groups, there was an inverse significant correlation between plasma BD-tau levels and Glasgow Outcome Scale – Extended (GOS-E) scores (spearman rho = -0.6727, p<0.0001), showing that higher BD-tau levels correspond with worse outcomes. In addition, there was a significant difference in the GOS-E score between those with sTBI and those with cmTBI [Supplemental Figure 4]. Moreover, when looking at the cohort, a significant positive correlation was seen between plasma and CSF BD-tau readings (r=0.6150, p=0.0039).

Supplemental Table 7 summarizes plasma/CSF BD-tau correlations with p-tau217, NfL, and GFAP, showing strong significant positive correlations in both matrices. In the sTBI group, BD-tau displayed a significant correlation with p-tau217 in both plasma (p=0.0005) and CSF (p=0.0002), but with the CSF correlation being understandably stronger than that of plasma (r=0.9667 and 0.5761, respectively). Similar BD-tau-p-tau217 correlations existed in the control and cmTBI groups. Plasma BD-tau was correlated with plasma NfL (r=0.8910, p<0.0001) and GFAP (r=0.5424, p=0.0011) in the sTBI group, whereas correlations in the cmTBI and control groups did not reach significance.

## Discussion

We have presented an independent validation of the first commercially available BD-tau assay recently released by the biotech company Quanterix®. BD-tau is a novel blood biomarker to detect neurodegeneration/brain injury, with promising results published for different neurological conditions including AD, TBI [6,18,19,21] and stroke [28]. However, previous studies have used in-house research use only (RUO) assays, with no commercially available methods to provide widespread standardized access to this biomarker. In the current study, we evaluated the assay’s robustness and precision, spike recovery, dilution linearity, and sample stability. We also assessed the lot-to-lot comparisons, and correlations in different matrices than plasma including serum and CSF. Finally, we showed the utility of BD-tau in a clinical TBI cohort. These findings show that the BD-tau assay fulfills key analytical parameters recommended by regulatory agencies and neurochemistry expert groups.

The high robustness and precision (based on CVs significantly lower than 20%) show that the assay test-retest variations are within limits allowable in clinical laboratories within and between runs [24].

Our results show that plasma samples with high concentrations can be diluted up to 16- fold (combination of on-board and bench dilutions) and still give reliable concentrations within the linear range of the calibration curve, with reproducible readings. Further dilution is not recommended as it may lead to inaccurate signals. Moreover, assay recovery was high; the concentrations of recombinant tau exogenously added to QC plasma samples were recovered with 86-96% accuracy.

Additionally, the assay demonstrated selectivity to recombinant 2N4R tau441 vs. big tau when titrated in buffer or in pooled plasma. The signals of samples spiked with various concentrations of recombinant peripheral-tau did not differ from the non-spiked samples, both of which were significantly lower than the corresponding signals for the 2N4R tau441 isoform which increased proportionally to the spike level.

Our sample stability results showed that as the number of freeze/thaw cycles increases, the levels of plasma BD-tau tend to increase, with significant increases at 6 and 8 f/t cycles.

This may be due to the disruption of protein complexes that lead to denaturation and release of free peptides. For storage temperature, our results show that storing samples at room temperature, 4°C, and -20°C for various periods of time does not significantly impact the levels of plasma BD-tau in any manner.

We also tested correlations of BD-tau levels in different matrices. Plasma-serum and CSF-plasma BD-tau pairs demonstrated strong correlations, indicating reliability independent of matrix choice in alignment with findings from previous studies [6,18,19]. Yet, the absolute concentrations in serum were significantly higher than those in paired plasma samples, which contradicts previous findings from an initial assay prototype [18]. In the TBI cohort, group differences in CSF and plasma were similar, meaning either matrix can be used but should be used independently in a whole study.

It is noteworthy that, despite being within the error-allowable margin for the tested QC samples, an unexplained apparent drift in signal was evident when the same samples were measured in wells at the start vs. at the end of the same plate in an identical run. This drift was observed when classifying assay signals using either AEBs values or estimated concentrations, indicating that this issue cannot be explained by potential standard curve signal differences between plates. Furthermore, our sample stability results showed that sample storage temperature (4°C or RT) did not impact BD-tau readings. Thus, such drift could not be due to temperature fluctuations between/during samples preparation and measurement. Additionally, consistency of the observation across different Simoa HD-X instruments rules out an instrument-related problem. Finally, paired t-testing on the number of beads for the same QC samples from the start and end of plates did not reveal statistical differences (p=0.1217), indicating the drift is likely not attributable to a difference in the amount of capture antibody.

We extended our analytical validation to clinical validation, examining a pilot TBI cohort. Our results demonstrated the utility of BD-tau in distinguishing individuals with severe traumatic brain injury at four days post-injury from those with chronic TBI, with only plasma BD-tau distinguishing severely injured patients from controls. This may be due to the small sample sizes in CSF groups. Moreover, plasma and CSF BD-tau were correlated, in agreement with previous findings [6,19,22]. Since BD-tau separated outcome scores of those with chronic from severe acute brain trauma in both plasma and CSF, BD-tau readings could facilitate predicting unfavorable outcome and overall clinical management in the acute stages of traumatic brain injury.

The correlation between plasma BD-tau with NfL and GFAP supports findings in other studies that BD-tau elevation in TBI can be an indicator of neurodegeneration/neuronal injury including in response to various processes including microglia activation [6,18,19,21,29]. A contradictory finding with the literature is the lack of CSF BD-tau correlation with NfL and GFAP for cmTBI or sTBI [22], perhaps due to the small CSF sample sizes, since they correlate in the full cohort. Perhaps the most intriguing correlation revealed in this study is the significant correlation between plasma BD-tau and p-tau217 across the full cohort and the sTBI and control sub-groups. P-tau217 is one of the most effective predictors of AD and related cognitive decline [7,30,31]. Larger cohorts are needed to verify these results. Another possibility is combining these biomarker measurements to help determine whether underlying neurodegeneration is due to AD when evaluating neurodegenerative diseases [7,21,30–32].

## Limitations

Although comprehensive, our study has limitations including our preliminary TBI cohort was relatively small and contained small within-diagnostic group numbers. In addition, the CSF samples were limited, precluding a direct comparison of findings in paired CSF and plasma samples from the same individuals.

## Conclusions

In summary, our study provides independent evidence that the plasma BD-tau assay kit has acceptable analytical performance as well as utility for evaluation of clinical TBI. Our findings have multiple future implications, including providing the basis for establishing guidelines for shipping and storage of plasma samples intended to test BD-tau levels. In addition to analytical findings, our results show that plasma BD-tau levels distinguish between severe TBI and chronic-mixed TBI, the former of which is associated with worse clinical outcomes as indicated by GOS-E scores. Thus, our results indicate that immediate BD-tau readings post-injury may provide an insight into the outcome and therefore allow for the formulation of an appropriate treatment plan.

## Data Availability

De-identified data generated in this study can be shared with qualified and identifiable investigators for the purpose of replicating the results and procedures in the study. Requests can be made to the corresponding author of the present study (T.K.K.) who will refer them to the respective cohort principal investigators where necessary. Requests will be reviewed by the investigators and respective institutions to ensure that each data request and transfer are in agreement with US legislation on general data protection or is subject to any intellectual property or confidentiality obligations. The purpose of these procedures is to ensure participant anonymity and ensure data safeguarding limited to the terms set forth in the IRB approvals. Data request can be made directly at https://www.adrc.pitt.edu/for-researchers/adrc-data-resources/ for Pittsburgh ADRC.

## Abbreviations

CTE: Chronic traumatic encephalopathy
CV: Coefficient of variation
CVr: Repeatability coefficient of variation
CVRw: Intermediate precision coefficient of variation
(U/L)LOQ: (Upper/Lower) limit of quantification
4-PL: Four-parameter logistic regression
AD: Alzheimer’s Disease
AD8: Ascertain Demential 8 item Questionnaire
ADRC: Alzheimer’s Disease Research Center
AEB: Average enzyme per bead
APOE: Apolipoprotein E
Aß: Amyloid beta
BD-tau Adv PLUS: BD-tau Advantage PLUS Kit
BD-tau: Brain-derived tau
cmTBI: Chronic-mixed TBI
CNS: Central nervous syste
CSF: Cerebrospinal fluid
CT: computed tomography
EDTA: ethylenediaminetetraacetic acid
F/T: freeze/thaw
FDA: Food and Drug Administration
GCS: Glasgow Coma Scale
GFAP: Glial-fibrillary acidic protein
GOS-E: Glasgow Outcome Scale - Extended Version
HD-X: Quanterix®’s trademark name for their instrument
NfL: neurofilament light chain
p-tau: phosphorylated tau
QC: Quality control
Simoa: Single-molecule array
sTBI: Severe-acute TBI
TBI: Traumatic brain injury
t-tau: Total tau
UCH-L1: Ubiquitin C-terminal hydrolase-L1.

### Human Genes

*APOE:* Apolipoprotein E

## Declarations

### Ethics approval and consent to participate

All patient samples were obtained with full written informed consent and approved by the University of Pittsburgh Institutional Review Board (ADRC study: MOD19110245-023; BTRC study: 19030228; Military CTE study: 19110161)

### Consent for publication

Not applicable.

## Authors’ contributions

TKK, MNN, and XZ contributed to the study’s conception and design. MNN ran biochemical assays on TBI and ADRC cohorts, performed data analysis and produced the figures. ADC, AMP, DOO, JKK, SES, OLL, XZ, and YC provided review and feedback on manuscript contents. YC provided plasma/serum ADRC samples used in matrix comparisons. AMP, DOO, and SES provided samples from TBI and BTRC cohort used in clinical validation. SES contributed to writing the methods and provided participant information for the TBI and BTRC cohorts. All authors contributed to and approved the final version of the manuscript.

## Acknowledgements

We express our appreciation to the Karikari Laboratory members, our collaborators at the Pitt-ADRC and the Puccio Laboratory.

