## Supplemental Appendix for "Plasma brain-derived tau: analytical and clinical validation of the first commercial immunoassay"

**1. Supplemental Methods**

#### 1.1 Analytical validation of the BD-tau Advantage PLUS assay

The analytical validation of the assay evaluated the assay’s robustness, precision, lower limit of quantification (LLOQ), specificity, dilution linearity, spike recovery, plasma/serum correlations, sample stability, and lot-to-lot variation.

##### *1.1.1 Robustness*

Internal quality control samples at low, medium, and high analyte levels were run 20 analytical times across 10 plates. Within plate robustness was determined by combining the coefficient of variation (CV) for each individual plate, and between plate robustness was evaluated by determining the CV across all samples.

##### *1.1.2 Precision*

Repeatability and intermediate precision were evaluated by calculating the coefficients of variation that were produced by running five analytical replicates of five different plasma samples, one time each day for five different days under the same conditions. Concentrations were then normalized to the average of all 25 measurements of the respective sample and portrayed as percentages. Repeatability (CV_r_) and intermediate precision (CV_Rw_) were then determined to assess within plate and inter-plate variation.

##### *1.1.3 Lower limit of quantification (LLOQ) determination*

Sixteen analytical runs of the blank (BD-tau Advantage PLUS calibrator diluent) were processed on the instrument. The average and standard deviation of the neat signals were calculated. The LLOQ was determined by using the inverse 4-parameter logistic (4PL) regression formula (Equation 1), with the desired signal being equal to the average of the 16 measured blanks added to 10 standard deviations.

Equation 1: $Concentration=C\left( \frac{A-D}{Signal-D}-1 \right)^{\frac{1}{B}}$

##### *1.1.4 Specificity*

Assay specificity was assessed by spiking four-fold diluted plasma samples and calibrator diluent with three different concentrations of either the CNS-abundant 2N4R-tau (BD-tau) or the peripherally enriched high molecular weight tau (Big-tau). The three concentrations were 100 pg/mL, 50 pg/mL, and 25 pg/mL, which spanned the dynamic range of the calibration curve. Non-spiked samples were also measured for reference.

##### *1.1.5 Dilution Linearity*

Three plasma samples were diluted four-fold with BD-tau+ plasma sample diluent and divided into seven aliquots. Calibrator stock solution was prepared by adding 1465 µL of calibrator diluent to lyophilized calibrator stock to create a 750 pg/mL stock solution. The stock was first diluted two-fold in the first plasma aliquot to 375 pg/mL, followed by six four-fold serial dilutions in diluted plasma to get the theoretical concentration below the analytical LLOQ, resulting in the following concentrations (pg/mL): 93.75, 23.44, 5.859, 1.465, 0.3662, and 0.0916. To assess relative error, concentrations were normalized to the average measurement of the undiluted sample and then adjusted for the fold-dilution by multiplying to the dilution factor. 80-120% after adjustment was considered to be linear dilution. R² values were calculated for non-diluted, four- and 16-fold diluted samples to portray linearity.

##### *1.1.6 Spike Recovery*

Two plasma samples were diluted four-fold with BD-tau Advantage PLUS plasma sample diluent and spiked with 25 µL of BD-tau calibrator stock solutions to four spike levels: 192 pg/mL, 96 pg/mL, 48 pg/mL, and 24 pg/mL. The assay buffer was also spiked with the same spikes as the samples to measure spike concentration in buffer, and the plasma sample was also run non-spiked to measure the neat concentration. Equation 2 was used to calculate the recovery at each spike level.

Equation 2: $\% Recovery= \frac{Concentration of spiked sample}{Concentration of non-spiked sample+Concentration of spike in diluent}$

##### *1.1.7 Plasma/Serum comparisons*

Fifty pairs of plasma/serum samples from a cohort from the University of Pittsburgh Alzheimer's Disease Research Center (Pitt-ADRC) were processed and concentrations values were compared. Pearson correlations between plasma and serum BD-tau readings were evaluated, and absolute plasma/serum concentrations were compared. Only pairs with values for both plasma and serum (n=48) were included in the analyses.

##### *1.1.8 Sample Stability*

Three plasma samples were divided into 19 aliquots containing an equal volume, then subjected to different conditions to test the impact of freeze/thaw (f/t) cycles and various storage temperatures/times on the level of analyte. A detailed description of conditions is provided in Supplemental Table 1. For each f/t cycle, samples were placed at room temperature for 2 hours, after at least 12 hours of being at -80°C.

### **2. Supplemental Figures**

#### Figure S1. Example calibration curve

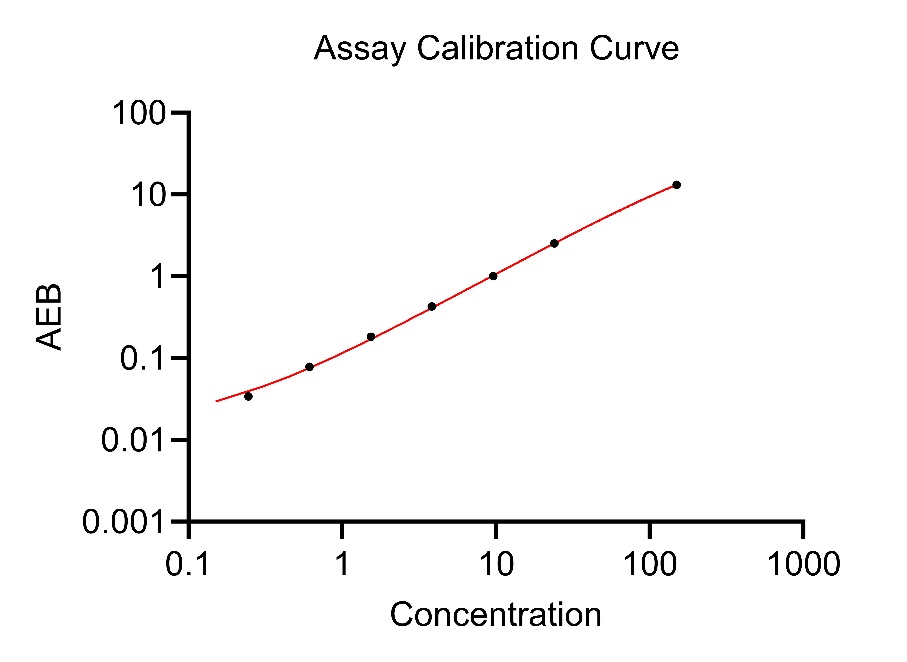

Example of a four-parameter logistic (4PL) regression calibration curve on a log-log scale. The curve was generated with seven non-zero calibrators that are used with the BD-tau Advantage PLUS assay.

#### Figure S2. Storage stability sample results

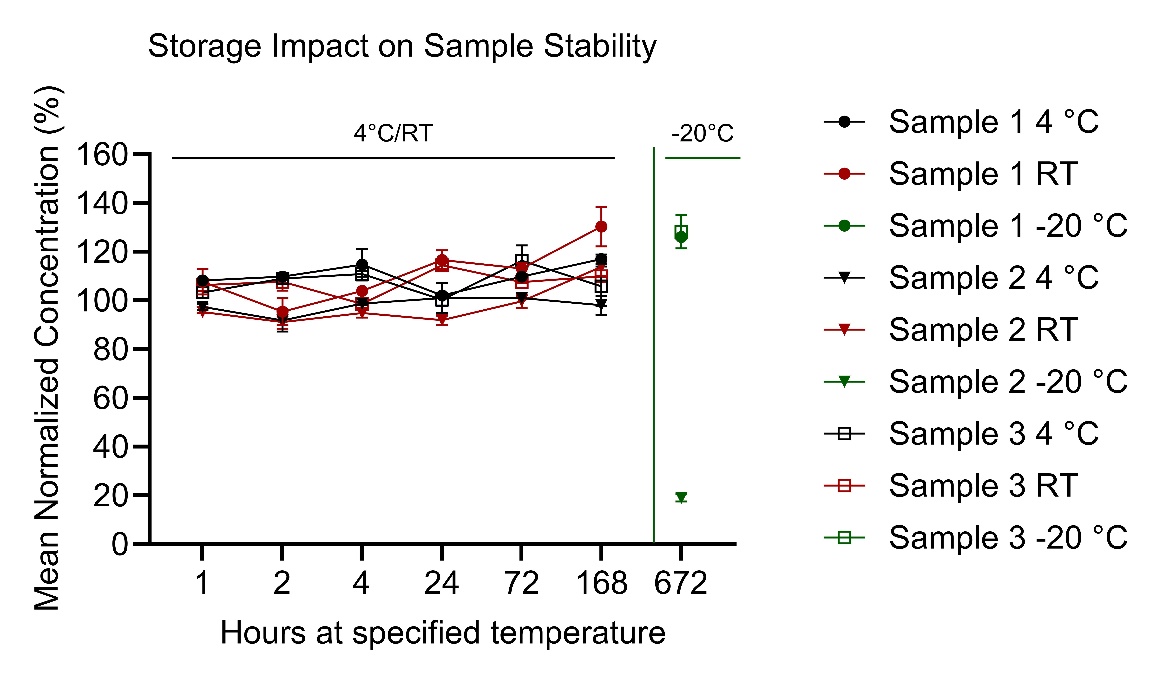

Normalized plasma BD-tau concentrations for three different samples initially stored at 4°C, room temperature (RT), or -20°C for various times before moving to -80°C.

#### Figure S3. Freeze/thaw sample stability results

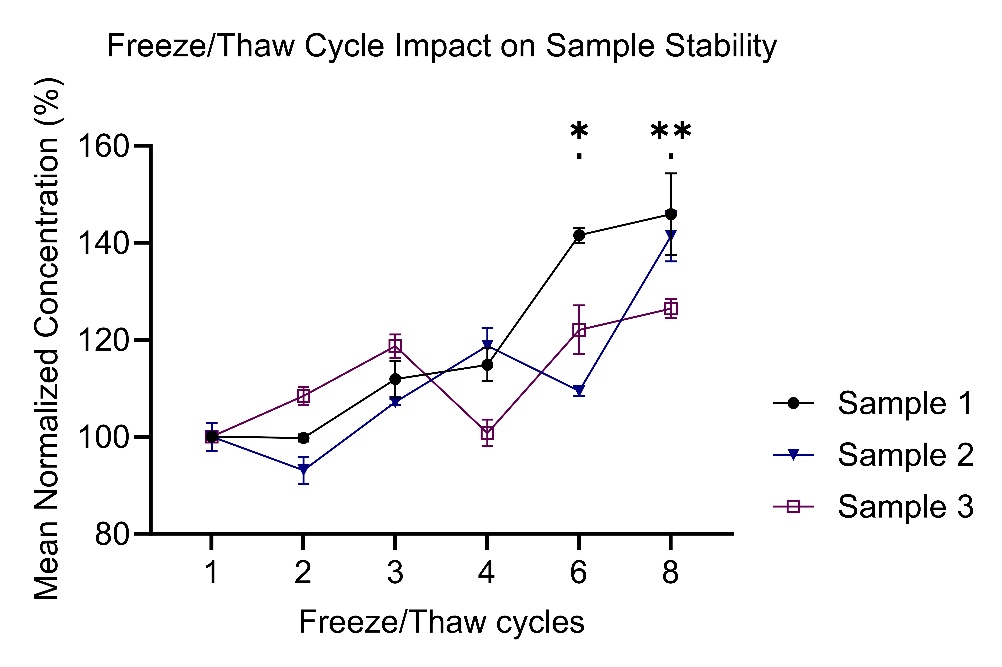

Impact of free/thaw (f/t) cycles on plasma BD-tau levels for three different plasma samples under the same f/t conditions. Significant differences from Dunn’s multiple comparisons test are portrayed by an asterisk above the cycles where significant differences from the reference sample (f/t = 1) were seen (*: p<0.05; **: p<0.01).

#### Figure S4. Glasgow Outcome Scale – Extended (GOS-E) score for severe and chronic mixed TBI

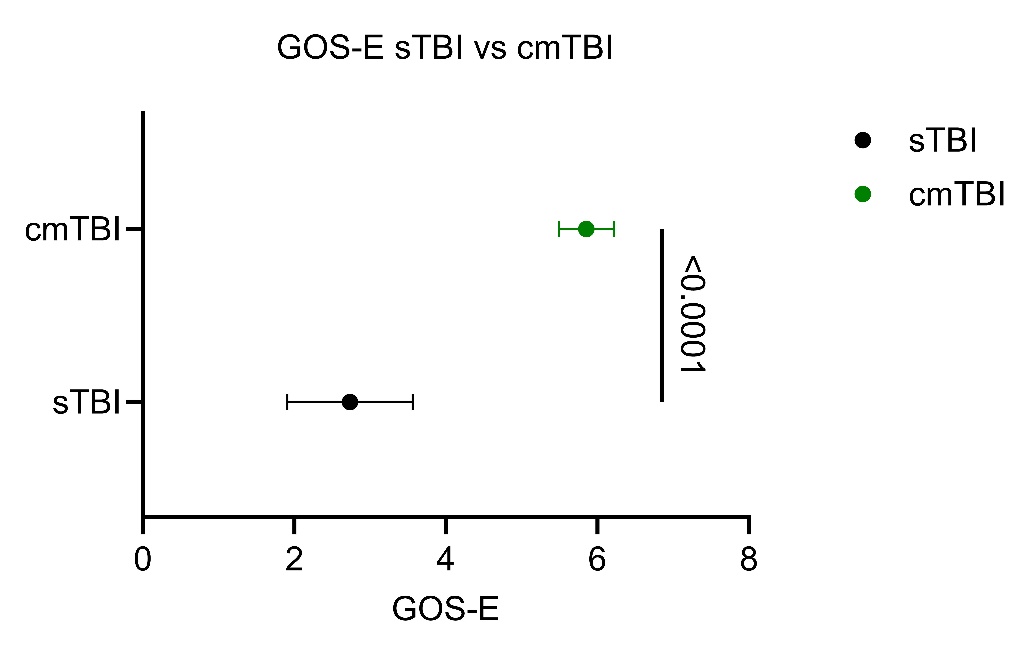

Forest plot showing the comparison of the Glasgow Outcome Scores – Extended version (GOS-E) for patients with severe acute traumatic brain injury (sTBI; n=34) and chronic mixed TBI (cmTBI; n=21). The mean ± 95% CI are reported.

### **3. Supplemental Tables**

#### Table S1. Sample stability storage conditions

| Aliquot | Initial storage | Time until -80ºC | Freeze/Thaw cycles |
| --- | --- | --- | --- |
| 1 | -80ºC |  | 1 |
| 2 | -80ºC |  | 2 |
| 3 | -80ºC |  | 3 |
| 4 | -80ºC |  | 4 |
| 5 | -80ºC |  | 6 |
| 6 | -80ºC |  | 8 |
| 7 | RT | 1 h |  |
| 8 | RT | 2 h |  |
| 9 | RT | 4 h |  |
| 10 | RT | 24 h |  |
| 11 | RT | 72 h |  |
| 12 | RT | 168 h |  |
| 13 | 4ºC | 1 h |  |
| 14 | 4ºC | 2 h |  |
| 15 | 4ºC | 4 h |  |
| 16 | 4ºC | 24 h |  |
| 17 | 4ºC | 72 h |  |
| 18 | 4ºC | 168 h |  |
| 19 | -20ºC | 28 days |  |

Description of freeze/thaw and storage conditions for plasma samples used in sample stability experiments. (h): hours.

#### Table S2. Demographics of pilot TBI cohort

|  | Control | Chronic-Mixed | Severe-Acute |
| --- | --- | --- | --- |
| **n** | 8 | 21 | 34 |
| **Age (mean ± SD)** | 38.4 ± 6.5 years | 40.4 ± 8.5 years | 53.0 ± 15.6 years |
| **Sex (n, % male)** | 7 (87.5%) | 17 (80.1%) | 27 (79.71%) |
| **Injury Type (#, %)** |  |  |  |
| **No Injury** | 3 (37.5%) | 0 (0%) | 0 (0%) |
| **Blunt** | 5 (62.5%) | 14 (66.7%) | 34 (100%) |
| **Blunt&Blast** | 0 (0%) | 7 (33.3%) | 0 (0%) |
| **Time from Injury (mean ± SD)** | 29.6 ± 5.5 years | 10.6 ± 6.7 years | 4 days |
| **Number of TBI (median, range)** | 1 (0-3) | 4 (2-14) | 1 |
| **GOS-E (median, range)** | n/a | 6 (5-8) | 1 (1-8) |

Traumatic brain injury (TBI) cohort patient demographics. Standard Deviation (SD); Glasgow Outcome Scale Extended (GOS-E). The controls were more than a year from their TBI and did not have any residual symptoms.

#### Table S3. Repeatability and intermediate precision

| Sample | Median Concentration (pg/mL) | %CV_r_ | %CV_Rw_ |
| --- | --- | --- | --- |
| Sample 1 | 8.53 | 10.2 | 12.0 |
| Sample 2 | 5.50 | 9.07 | 9.95 |
| Sample 3 | 7.97 | 12.4 | 15.1 |
| Sample 4 | 6.32 | 6.47 | 9.52 |
| Sample 5 | 6.46 | 11.7 | 11.7 |

Median concentrations, and repeatability (CV_r_) and intermediate precision (CV_RW_) coefficients of variation for plasma samples in precision tests.

#### Table S4. LLOQ AEB values

| Blank | AEB |
| --- | --- |
| 1 | 0.00874 |
| 2 | 0.00580 |
| 3 | 0.00581 |
| 4 | 0.00633 |
| 5 | 0.00807 |
| 6 | 0.00510 |
| 7 | 0.00543 |
| 8 | 0.00561 |
| 9 | 0.00654 |
| 10 | 0.00429 |
| 11 | 0.00543 |
| 12 | 0.00571 |
| 13 | 0.00562 |
| 14 | 0.00509 |
| 15 | 0.00567 |
| 16 | 0.00657 |

AEB values for calibrator diluent samples used in the lower limit of quantification (LLOQ) determination.

#### Table S5. Specificity tests AEB values

| Spike  concentration  (pg/mL) | Median AEB | | | |
| --- | --- | --- | --- | --- |
|  | **BD-tau with assay buffer** | **Big-tau with assay buffer** | **BD-tau with 4x QCD** | **Big-tau with 4x QCD** |
| 0 | 0.01 | 0.01 | 0.13 | 0.13 |
| 25 | 5.86 | 0.03 | 4.88 | 0.14 |
| 50 | 11.7 | 0.02 | 8.73 | 0.14 |
| 100 | 20.3 | 0.02 | 15.6 | 0.14 |

Median average enzyme per bead (AEB) values for specificity samples in both diluent and four-fold diluted QC plasma.

#### Table S6. Spike recovery results

| Spike | Concentration In Buffer | Sample 1 Concentration | % Recovery | Sample 2 Concentration | % Recovery |
| --- | --- | --- | --- | --- | --- |
| 0 | N/A | 1.19 | N/A | 1.67 | N/A |
| 24.0 | 22.5 | 22.8 | 96.0 | 21.7 | 89.9 |
| 48.0 | 45.4 | 41.8 | 89.9 | 40.5 | 86.0 |
| 96.0 | 87.7 | 82.6 | 92.9 | 79.6 | 89.0 |
| 192 | 177 | 158 | 88.5 | 160 | 89.2 |

Spike recovery results explanation showing the expected spike concentration in buffer, observed spike concentration in buffer, the observed concentrations of samples before spiking, and the observed concentrations and percentage recovery of samples spiked at four different calibrator stock concentrations.

#### Table S7. Biomarker correlations in TBI

|  |  | BD-tau vs p-tau217 | | | BD-tau vs NfL | | | BD-tau vs GFAP | | |
| --- | --- | --- | --- | --- | --- | --- | --- | --- | --- | --- |
|  | **Group** | **N** | **r** | **P-value** | **N** | **r** | **P-value** | **N** | **r** | **P-value** |
| Plasma | Control | 8 | 0.857 | 0.0107 | 8 | 0.333 | 0.428 | 8 | 0.571 | 0.151 |
|  | cmTBI | 21 | 0.132 | 0.569 | 21 | -0.111 | 0.632 | 21 | -0.227 | 0.323 |
|  | sTBI | 33 | 0.576 | 0.0005 | 33 | 0.891 | <0.0001 | 33 | 0.542 | 0.0011 |
|  | Entire Cohort | 63 | 0.742 | <0.0001 | 63 | 0.880 | <0.0001 | 63 | 0.819 | <0.0001 |
| CSF | Entire Cohort | 20 | 0.773 | <0.0001 | 18 | 0.730 | 0.0006 | 18 | 0.707 | 0.0010 |

Spearman correlations between BD-tau and each of p-tau217, NfL, and GFAP in the entire TBI cohort and its subgroups (controls, cmTBI, and sTBI) in plasma. Spearman correlations between BD-tau and the other biomarkers in CSF are only shown for the entire cohort due to the small sample size of the subgroups.
